# Associations with SARS-CoV-2 Positivity Among People in Refugee Settlements: An Interactive Voice Response Survey in Uganda

**DOI:** 10.64898/2026.10.20.26365986

**Authors:** Timothy R. Muwonge, Robin E. Klabbers, Scovia Ajidiru, Phuong Pham, Patrick Vinck, Andrew Mujugira, Rogers Nsubuga, Monisha Sharma, Sukanya Borthakur, Numan Mohammed, R Parkes-Ratanshi, Connie Celum, Kelli N. O’Laughlin

**Author notes:** **Corresponding author**, Timothy R Muwonge, MBChB, MPH, Research Department, Infectious Diseases Institute, Makerere University, Kampala, Uganda. **Data Availability** Due to the sensitive nature of demographic and geographic information related to refugee populations, the dataset generated and analyzed during this study is not publicly available. However, de-identified data may be shared with academic researchers upon reasonable request. Each request will be reviewed on a case-by-case basis to ensure alignment with ethical standards and to safeguard the confidentiality and safety of study participants. Requests for data access should be directed to the Makerere University School of Health Sciences Research Ethics Committee, chaired by Dr. Kalidi Rajat. The Secretariat contact is Stella Imot.

## Abstract

**Introduction:** Limited SARS-CoV-2 testing capacity during the early COVID-19 pandemic required approaches to identify individuals at highest risk of infection for prioritized testing. We assessed the association between symptoms and exposures reported at the time of SARS-CoV-2 testing and test positivity among individuals referred for testing through an interactive voice response (IVR) platform in refugee settlements in Uganda.

**Methods:** We used Dial-COVID, a multilingual, toll-free interactive voice response (IVR) platform to survey residents of refugee settlements in Uganda regarding COVID-19 symptoms, exposures, and comorbidities. Respondents reporting symptoms or exposures were invited for SARS-CoV-2 testing using nasal or oropharyngeal swab specimens. At the time of testing, an interviewer-administered questionnaire assessed symptoms and exposures occurring within the preceding two weeks. Log-binomial regression was used to identify factors associated with SARS-CoV-2 positivity.

**Results:** Between February 2021 and March 2022, 6,913 of 15,467 Dial-COVID callers (44.7%) completed the IVR survey. Among survey respondents, 5,184 (75.0%) reported at least one symptom or exposure. Of these, 1,361 (26.3%) were invited for testing, and samples were collected from 751 (55.2%) within seven days of survey completion. SARS-CoV-2 test results were available for 741 individuals, of whom 48 tested positive (6.5%). Among symptoms reported at the testing visit, severe muscle pain (risk ratio [RR] 2.08; 95% CI 1.20–3.57) and unusual fatigue (RR 2.34; 95% CI 1.36–4.03) were associated with SARS-CoV-2 positivity in unadjusted analyses. After adjustment for sex, refugee status, hypertension, and diabetes, only unusual fatigue remained independently associated with a positive test result (aRR 2.10, 95% CI 1.20– 3.67).

**Conclusion:** Few self-reported symptoms were associated with SARS-CoV-2 positivity, highlighting the challenges of prioritizing testing based on symptoms alone. IVR-based platforms may provide a feasible approach for screening, surveillance, and linkage to testing in resource-constrained humanitarian settings.

## Introduction

At the onset of the Coronavirus Disease 2019 (COVID-19) pandemic, refugee settlements were considered particularly vulnerable to SARS-CoV-2 outbreaks because of high population density, limited healthcare resources, and barriers to care (1, 2). Limited testing capacity during the early phases of the pandemic complicated efforts to identify and respond to cases in humanitarian settings, creating a need for approaches to identify individuals at highest risk of infection for prioritized testing and surveillance.

Internet-based symptom trackers had been used to map the spread of COVID-19, inform outbreak projection models, and target resources and public health messaging (1–4), but were considered less feasible in refugee settlements, where smartphone ownership and internet connectivity are limited. Interactive voice response (IVR) technology, accessible via basic mobile phones, offered a potentially scalable alternative for symptom surveillance and public health outreach in such settings. Prior studies have demonstrated the feasibility of IVR-based data collection in refugee populations (5) and its successful use in Uganda to support HIV treatment adherence among adults and adolescents (6, 7).

Few studies have evaluated the use of IVR platforms in humanitarian settings during the COVID-19 pandemic. We used Dial-COVID, a multilingual IVR platform, to identify individuals living in refugee settlements in Uganda reporting COVID-19 symptoms or exposure for SARS-CoV-2 testing. Symptom-based prioritization is only useful if reported symptoms are associated with infection. We therefore assessed the association between symptoms and exposures reported at the time of testing and SARS-CoV-2 positivity.

## Methods

### Dial-COVID study design

The Dial-COVID study was a cross-sectional study conducted through a collaboration between academic institutions in Uganda (Infectious Diseases Institute [IDI], Makerere University) and the United States (the University of Washington, Massachusetts General Hospital, and Brigham Women’s Hospital), the non-governmental organization Medical Teams International (MTI), and the telecommunications company Viamo, as previously reported (8). The Dial-COVID IVR tool was implemented to collect individual-level data on COVID-19 symptoms and exposures and disseminate COVID-19-related information in refugee settlements in Uganda. From February 2021 through March 2022, the Dial-COVID platform was advertised in refugee settlements in Uganda (Adjumani, Kyaka II, Kyangwali, Nakivale, Oruchinga, Palorinya, and Rwamwanja Refugee Settlement) **(Figure 1)**. Individuals with symptoms or known exposure, or who were interested in learning more about COVID-19 were encouraged to “beep” the Dial-COVID platform, i.e, place a call to the Dial-COVID platform and hang up, which prompted a free return call from the platform. In the return call, participants received the Dial-COVID symptom survey, a set of pre-recorded IVR questions about COVID-19 symptoms and exposures (8) (APPENDIX 1). Participants responded to survey questions by selecting the corresponding number on their mobile phone keypad. Based on their answers, participants received one of two Ugandan Ministry of Health guidelines aligned with public health messages tailored to their risk profile (high-risk vs low-risk).

**Figure 1.**
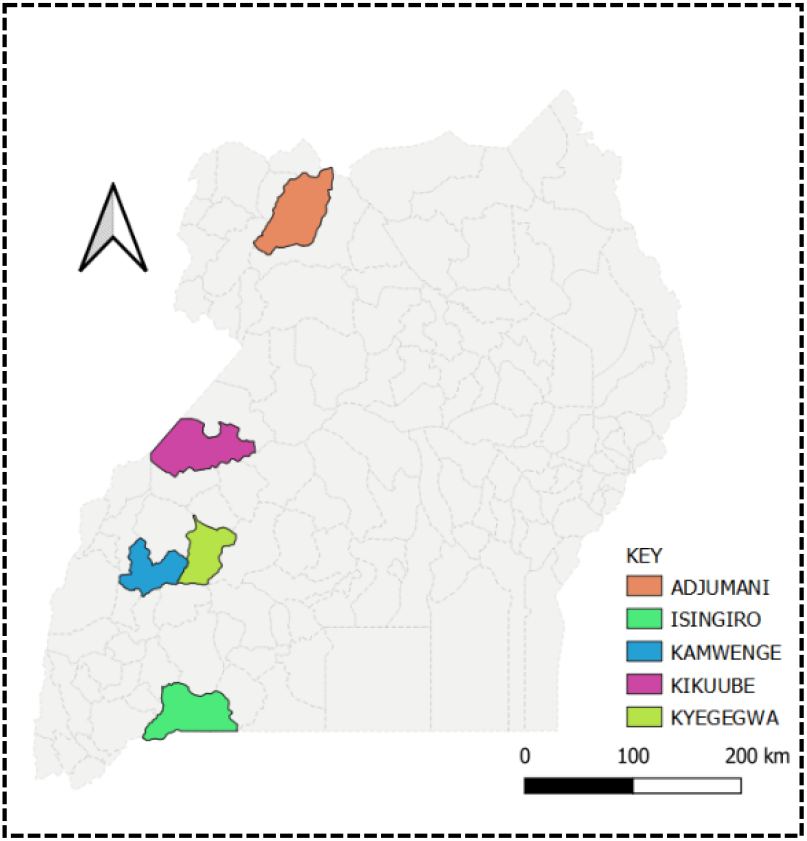
Refugee Settlement Districts

### Subject selection and data collection

Dial-COVID call-in participants residing in refugee settlements who reported at least one COVID-19 symptom or exposure in the Dial-COVID IVR survey and who indicated a willingness to participate in additional COVID-19-related research were identified. A convenience sample of those participants located near a health center accessible to study staff within 7 days of Dial-COVID survey completion were invited by phone to come for SARS-CoV-2 testing (Fig 1). Research Assistants, over the phone and in person, ensured that only Dial-COVID survey participants were invited by first confirming participants’ identities by inquiring about their prior participation in Dial-COVID, their reported age, sex, and general location of residence.

### Informed Consent

A research assistant (RA) obtained and documented verbal informed consent for testing in the participant’s study language of choice (Arabic, Dinka, English, Kakwa, Kinyarwanda, Kiswahili, Lugbara, Runyankore, and Somali), with an interpreter as needed. The consent process was conducted either in person, following COVID-19 safety measures such as wearing personal protective equipment and maintaining appropriate physical distance, or by phone to minimize COVID-19 exposure and transmission risks, based on participant preference. During the consent process, participants were informed that their test results would be shared with the Ugandan Ministry of Health Task Force, a team responsible for notifying individuals with a positive SARS-CoV-2 test result, linking them to a treatment center if necessary, and conducting contact tracing.

### Intake survey

At the testing visit, which occurred an average of 7 days after completion of the Dial-COVID survey, following informed consent, the RA administered an intake survey either to collect demographic information (age, sex, refugee status, marital status, religion, and country of origin), food security, health status (pre-existing comorbidities, pregnancy, general health), healthcare access, and SARS-CoV-2 exposure risk (work at a health facility, visiting public/crowded places, frequency of face mask use). Additionally, participants were asked whether they had experienced any of the following symptoms during the preceding 2 weeks: 1) cold symptoms (cough, fever, shortness of breath or otherwise feeling sick); 2) feeling feverish or having chills; 3) new or worsening cough; 4) persistent cough, (coughing a lot for more than an hour; or three or more episodes in 24 hours); 5) shortness of breath or chest pain/chest tightness; 6) loss of ability to smell or taste; 7) unusually strong muscle pains; 8) a sore throat or an unusually hoarse voice; 9) unusual fatigue; and 10) feeling tired and or struggling to get out of bed (see survey Appendix 1).

### SARS-CoV-2 testing

Laboratory technicians collected a nasal or oropharyngeal swab sample from the participants. Samples were placed in BD universal viral transport medium-embedded biohazard courier containers with dry ice before being transported by designated national couriers to the IDI Core Laboratory at Mulago Hospital in Kampala, where they were tested for SARS-CoV-2 using polymerase chain reaction methods within 48 hours (9). The Core Laboratory emailed test results to both the Ugandan Ministry of Health Task Force, which notified participants on behalf of the government, and study research assistants who provided parallel phone notifications to participants within one week of testing on behalf of the study. When multiple attempts to reach a participant were unsuccessful, or someone else answered the phone, a generic text message without identifiable information was sent to the participant, indicating either “SARS-CoV-2 test positive” or “SARS-CoV-2 test negative”. Participants could collect their official SARS-CoV-2 test result certificate from the facility if they desired. RAs referred participants who tested positive to MTI Program Officers, who, depending on the disease severity, either provided information on home-based care or linked participants to care and treatment at the nearest referral facility.

### Data Analysis

Participant characteristics, including demographics, symptoms, and comorbidities, were summarized descriptively and compared by SARS-CoV-2 test result using chi-square tests for categorical variables and t-tests for continuous variables. Variables associated with SARS-CoV-2 positivity at p<0.10 in bivariate analyses were considered for multivariable modeling. The primary objective was to identify symptoms associated with SARS-CoV-2 positivity. Symptoms associated with SARS-CoV-2 positivity in bivariate analyses were evaluated in a multivariable model adjusted for demographic characteristics associated with SARS-CoV-2 positivity (sex and refugee status). An additional model further adjusted for comorbidities associated with SARS-CoV-2 positivity (hypertension and diabetes). A model with and without adjustment for comorbidities is presented because these conditions may be more strongly associated with COVID-19 severity than risk of infection. Since severe muscle pain and unusual fatigue were highly correlated, these symptoms were evaluated in separate multivariable models to avoid collinearity. As a sensitivity analysis, we fitted an additional multivariable model that included all demographic characteristics and symptoms simultaneously. Adjusted risk ratios (aRRs) and 95% confidence intervals (CIs) were estimated using log-binomial regression models with robust standard errors. Statistical significance was defined as p<0.05. All analyses were conducted using Stata version 18 (StataCorp, College Station, TX).

## Results

The Dial-COVID platform received 15,467 calls, of which 6,913 (44.7%) resulted in a completed IVR survey. Among callers who completed a survey, 5,184 (75.0%) reported at least one COVID-19 symptom or exposure. Of these callers, a convenience sample of 1,361 (26.3%) was invited for testing, and samples were collected from 751 (55.2%). SARS-CoV-2 test results were available for 741 individuals (results for 10 participants were unavailable due to misplaced samples), of whom 48 (6.5%) tested positive.

Among participants with available test results, the median age was 28 years (interquartile range [IQR] 24–31), and most were male (66.1%), refugees (97.8%), and married (72.2%) (Table 1). The most commonly reported symptoms within the previous two weeks were cold symptoms (96.5%) and fever or chills (89.8%). Comorbidities were uncommon, with hypertension (5.1%) and diabetes (2.4%) being the most frequently reported. In bivariate analyses, female sex, refugee status, hypertension, and diabetes met the p<0.10 threshold for inclusion in multivariable models.

**Table 1.** Characteristics of participants with documented SARS-CoV-2 test results.

| Individual Characteristics | Overall N=741 | Tested COVID+ N=48 | Tested COVID- N=693 | p-value |
| --- | --- | --- | --- | --- |
| Mean (SD) or N (%) |  |  |  |  |
| **Female** | 251 (33.9) | 11 (22.9) | 240 (34.9) | 0.09 |
| **Age (years)** | 28.3 (6.4) | 28.9 (6.6) | 28.2 (6.4) | 0.51 |
| **Refugee Status** |  |  |  | 0.09 |
| Refugee | 725 (97.8) | 46 (95.8) | 679 (98.0) |  |
| Ugandan IDP* | 12 (1.6) | 1 (2.1) | 11 (1.6) |  |
| Ugandan non-IDP* | 2 (0.3) | 1 (2.1) | 1 (0.1) |  |
| Non-Ugandan, non IDP* | 2 (0.3) | 2 (0.3) | 0 (0.0) |  |
| **Relationship Status** |  |  |  | 0.48 |
| Married/living together | 535 (72.2) | 40 (83.3) | 495 (71.4) |  |
| Divorced/separated/widowed | 66 (8.9) | 4 (8.3) | 62 (8.9) |  |
| Single | 134 (18.1) | 4 (8.3) | 130 (18.8) |  |
| Missing Status | 6 (0.8) | 0 (0.0) | 6 (0.9) |  |
| **Comorbidities** |  |  |  |  |
| Asthma | 34 (4.6) | 1 (2.1) | 33 (4.8) | 0.39 |
| Chronic Lung Disease | 30 (4.0) | 3 (6.3) | 27 (3.9) | 0.42 |
| Smoker | 21 (2.8) | 3 (6.3) | 18 (2.6) | 0.14 |
| Hypertension | 38 (5.1) | 8 (16.7) | 30 (4.3) | <0.01 |
| Heart Disease | 19 (2.6) | 2 (4.2) | 17 (2.5) | 0.48 |
| Diabetes | 18 (2.4) | 4 (8.3) | 14 (2.0) | 0.01 |
| HIV | 21 (2.8) | 1 (2.1) | 20 (2.9) | 0.75 |
| **Symptoms experienced in the last 2 weeks,** |  |  |  |  |
| Cold symptoms | 715 (96.5) | 47 (97.9) | 668 (96.4) | 0.58 |
| Feverish or chills | 662 (89.8) | 43 (89.6) | 619 (89.8) | 0.96 |
| New or worsening cough | 197 (26.8) | 10 (20.8) | 187 (27.2) | 0.33 |
| Shortness of breath | 187 (25.4) | 9 (19.2) | 178 (25.8) | 0.31 |
| Loss of smell or taste | 176 (23.9) | 13 (27.1) | 163 (23.7) | 0.59 |
| Severe Muscle Pain | 202 (27.3) | 21 (43.8) | 181 (26.2) | 0.01 |
| Sore Throat | 109 (14.7) | 7 (14.6) | 102 (14.7) | 0.98 |
| Unusual fatigue | 209 (28.2) | 23 (47.9) | 186 (26.9) | <0.01 |

\*Internally Displaced Persons (IDP)

Severe muscle pain and unusual fatigue were associated with SARS-CoV-2 positivity in unadjusted analyses (severe muscle pain: risk ratio [RR] 2.08, 95% CI 1.20–3.59; unusual fatigue: RR 2.34, 95% CI 1.36–4.03) (Table 2). Because these symptoms were highly correlated, with 86.6% of participants reporting one symptom also reporting the other, they were evaluated in separate multivariable models. After adjustment for sex and refugee status, both severe muscle pain (aRR 2.10, 95% CI 1.21–3.66) and unusual fatigue (adjusted risk ratio [aRR] 2.40, 95% CI 1.40–4.12) remained associated with SARS-CoV-2 positivity. After additional adjustment for hypertension and diabetes, only unusual fatigue remained associated with SARS-CoV-2 positivity (aRR 2.10, 95% CI 1.20–3.67), whereas the association with severe muscle pain was attenuated (aRR 1.77, 95% CI 0.99–3.13).

**Table 2.** Symptom associations with SARS-CoV-2 Positivity (Relative Risks)

|  |  |  |  |
| --- | --- | --- | --- |
| **Symptoms** | **Unadjusted Model** | **Adjusted Model 1\*** | **Adjusted Model 2\*\*** |
| Severe Muscle Pain | RR=2.08 | RR=2.10 | RR=1.77 |
| 95%CI | (1.20, 3.59) | (1.21, 3.66) | (0.99, 3.13) |
| p-value | <0.01 | <0.01 | 0.05 |
| Unusual fatigue | RR=2.34 | RR=2.40 | RR=2.10 |
| 95%CI | (1.36, 4.03) | (1.40, 4.12) | (1.20, 3.67) |
| p-value | <0.01 | <0.01 | <0.01 |

\* Adjusted for sex and refugee status, \*\*Adjusted sex, refugee status, hypertension, and diabetes

In a sensitivity analysis including all demographic characteristics and symptoms in a single model, unusual fatigue remained independently associated with SARS-CoV-2 positivity (RR 2.51, 95% CI 1.13–5.57), whereas no other symptoms were significantly associated with infection (Table 3).

**Table 3.**
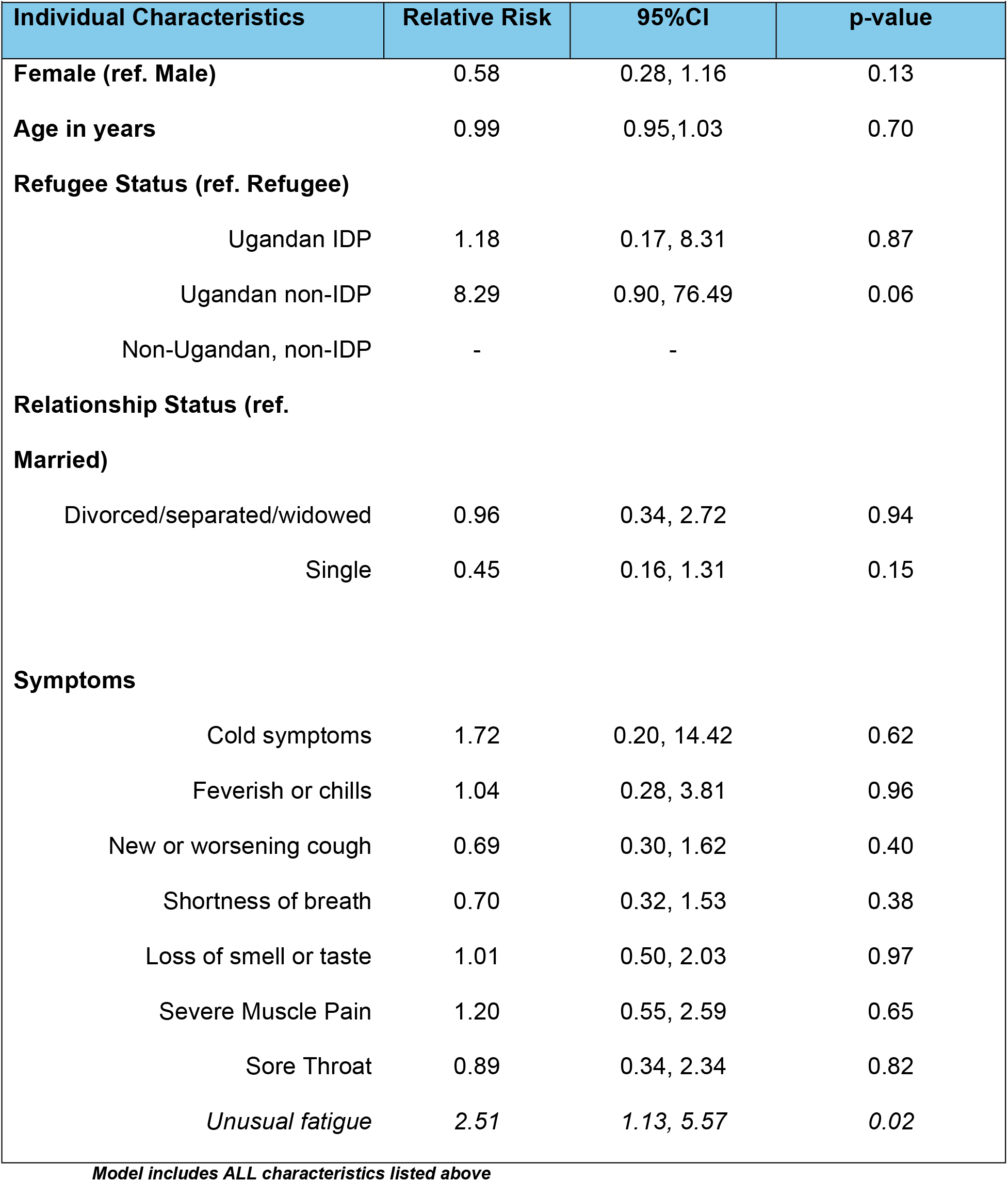
Demographic and Symptoms Associations withSARS-CoV-2 Positivity.

## Discussion

This interactive voice response study examined the association between self-reported symptoms and SARS-CoV-2 positivity among individuals referred for testing through the Dial-COVID IVR platform in refugee settlements in Uganda during a period of limited testing capacity. Few symptoms were associated with SARS-CoV-2 positivity, and unusual fatigue was the only symptom that remained independently associated with a positive test result after adjustment for demographic characteristics and comorbidities.

The limited predictive value of symptoms observed in this study is consistent with the broader literature on COVID-19 symptomatology. A Cochrane review of 44 studies involving more than 26,000 participants found that common COVID-19 symptoms generally have poor diagnostic accuracy (16). While symptoms such as cough and fever demonstrated relatively high sensitivity, their specificity was low, and fatigue showed only modest diagnostic performance. Similarly, although unusual fatigue was associated with SARS-CoV-2 positivity in our study, no symptom alone provided strong discriminatory value. Together, these findings highlight the challenges of prioritizing SARS-CoV-2 testing based solely on symptom profiles.

Although symptom-based prioritization had limited ability to distinguish individuals with SARS-CoV-2 infection from those without, the Dial-COVID platform demonstrated substantial reach, receiving more than 15,000 calls and facilitating nearly 7,000 completed symptom and exposure surveys. These findings suggest that the principal limitation was not the ability to collect symptom information at scale, but rather the non-specific clinical presentation of COVID-19 itself. IVR-based approaches may therefore have greater utility for surveillance and testing prioritization during outbreaks of diseases characterized by more distinctive symptom profiles. For example, Ebola virus disease, which is characterized by a more distinctive clinical presentation and well-defined epidemiologic risk factors than COVID-19, may be more amenable to symptom-based surveillance and testing prioritization. In addition, the platform’s ability to engage geographically dispersed and linguistically diverse populations highlights its potential value for public health communication, surveillance, and outbreak response in humanitarian settings.

Several limitations should be considered when interpreting these findings. First, because phones were frequently shared and only limited identifying information was collected via the IVR platform, IVR survey responses could not be directly linked to individual testing outcomes. As a result, symptom analyses were based on symptoms and exposures reported during the testing encounter rather than those reported through the IVR platform. Consequently, this study does not directly evaluate the predictive value of IVR-reported symptoms for SARS-CoV-2 infection. Second, participation in testing was limited to a convenience sample of callers who reported symptoms or exposures and were able to report for testing at selected health centers, which may have introduced selection bias. Third, symptoms were self-reported and referred to the preceding two weeks, creating the potential for recall bias. Fourth, the relatively small number of positive SARS-CoV-2 cases limited statistical power and the precision of effect estimates. Finally, the number of individuals who could be tested within a short period following IVR screening was constrained by geography and available resources, limiting our ability to evaluate the utility of the platform for identifying transmission hotspots.

## Conclusion

While self-reported symptoms had limited utility for identifying SARS-CoV-2 infection, the Dial-COVID study demonstrates that IVR technology can support large-scale collection of symptom and exposure information and facilitate linkage to testing among geographically dispersed, multilingual populations in humanitarian settings. IVR platforms may provide a practical approach for surveillance, risk communication, and outbreak response in resource-constrained settings.

## Data Availability

Due to the sensitive nature of demographic and geographic information related to refugee populations, the dataset generated and analyzed during this study is not publicly available. However, de-identified data may be shared with academic researchers upon reasonable request. Each request will be reviewed on a case-by-case basis to ensure alignment with ethical standards and to safeguard the confidentiality and safety of study participants. Requests for data access should be directed to the Makerere University School of Health Sciences Research Ethics Committee, chaired by Dr. Kalidi Rajat. The Secretariat contact is Stella Imot.

## Acknowledgements

We extend our sincere appreciation to the study research assistants and the Village Health Teams from Medical Teams International, whose unwavering commitment and hard work were instrumental to the success of this study. We are also grateful to collaborators from the United Nations High Commissioner for Refugees in Uganda and the Office of the Prime Minister for their invaluable support. Most importantly, we thank all the refugee and Ugandan participants for their generous participation.

